# Childhood Adversity and COVID-19 Outcomes in the UK Biobank

**DOI:** 10.1101/2023.03.20.23287479

**Authors:** Jamie L Hanson, Kristen O’Connor, Dorthea J Adkins, Isabella Kahhale

**Author notes:** Correspondence to: Jamie Hanson, Ph.D., *Learning Research & Development Center University of Pittsburgh, Murdoch Building 3420 Forbes Ave. Rm. 639 Pittsburgh, PA 15260. **Contributor Roles Taxonomy (CRediT) Author Statement:** Conceptualization: Hanson designed the study and developed the initial research question. Methodology: Hanson, O’Connor, Adkins, and Kahhale designed the study methodology and aided in identified relevant data collection instruments. Data curation: Hanson and Kahhale collected and organized the data. Formal analysis: Hanson conducted the statistical analysis and interpreted the results. Writing – original draft: Hanson drafted the manuscript. Writing – review & editing: O’Connor, Adkins, and Kahhale reviewed and edited the manuscript. Visualization: Hanson and Adkins created the figures and tables. Funding acquisition: Hanson secured funding for the project. **Declarations:** The corresponding author attests that all listed authors meet authorship criteria and that no others meeting the criteria have been omitted. **Competing interests:** All authors have completed the ICMJE uniform disclosure form at http://www.icmje.org/disclosure-of-interest/ and declare: no support from any organization for the submitted work; no financial relationships with any organizations that might have an interest in the submitted work in the previous three years; no other relationships or activities that could appear to have influenced the submitted work. **Funding:** This work was funded by internal grants provided to Dr. Hanson by the University of Pittsburgh.

## Abstract

**Objectives:** To investigate the association between childhood adversity and COVID-19-related hospitalization and COVID-19-related mortality in the UK Biobank.

**Design:** Cohort study.

**Setting:** United Kingdom.

**Participants:** 151,200 participants in the UK Biobank cohort who had completed the Childhood Trauma Screen, were alive at the start of the COVID-19 pandemic (01-10-2021), and were still active in the UK Biobank when hospitalization and mortality data were most recently updated (11-2021).

**Main outcome measures:** COVID-19-related hospitalization and COVID-19-related mortality.

**Results:** Higher self-reports of childhood adversity were related to greater likelihood of COVID-19-related hospitalization in all statistical models. In models adjusted for age, ethnicity, and sex, childhood adversity was associated with an OR of 1.227 of hospitalization (95% CI=1.153 to 1.306, Childhood Adversity *z*=6.49, *p*<0.005) and an OR of 1.25 of a COVID-19 related death (95% CI=1.11 to 1.424, Childhood Adversity *z*=3.5, *p*<0.005). Adjustment for potential confounds attenuated these associations, although associations remained statistically significant.

**Conclusions:** Childhood adversity was significantly associated with COVID-19-related hospitalization and COVID-19-related mortality after adjusting for sociodemographic and health confounders. Further research is needed to clarify the biological and psychosocial processes underlying these associations to inform public health intervention and prevention strategies to minimize COVID-19 disparities.

**Trial registration:** Work Completed under UK Biobank Project ID 92699 (“*Associations between COVID-19 Symptoms* & *Stressful Life Experiences*”).

**Summary Prompts:** *What is already known on this topic:*

- Disparities in COVID-19 outcomes are driven by numerous health and sociodemographic risk factors
- Childhood adversity is associated with lifelong physical health disparities and early mortality
- No known studies to date have examined the association between childhood adversity and COVID-19 mortality and morbidity

*What the study adds:*

- In the UK Biobank, childhood adversity was significantly associated with COVID-19-related hospitalization and COVID-19-related mortality.
- For both morbidity and mortality, these links were seen in statistical models adjusted for important sociodemographic and physical health confounders.

*How this study might affect research, practice or policy:*

- Modifiable and more proximal psychosocial factors may impact adult health outcomes, including COVID-19-related mortality and hospitalization
- Adversity may relate to depression, self-concept, or self-regulation, cascading from childhood experiences to the outcomes that we investigated here.
- Pinpointing these processes may allow for policy and interventions to lessen the negative impact of COVID-19 in those that have suffered childhood adversity.

## INTRODUCTION

Since 2020, COVID-19 has claimed the lives of over 6.7 million people globally [1]. This pandemic has led to unprecedented global declines in life expectancy, with COVID-19 being the leading cause of death in the Americas and the third leading cause of death in Europe [2,3]. COVID-19 has also placed enormous burdens on health care systems, often requiring hospitalization and intensive care for those with severe infections, in turn leading to billions of dollars in healthcare expenses [4]. While COVID-19 is a significant public health issue, the factors contributing to COVID-19 mortality and morbidity are still unclear. With this pandemic linked to significant long-term negative sequelae (e.g., changes in brain structure and risk for heart conditions) [5,6], it is critical to increase knowledge about factors contributing to risk in order to guide public health intervention and prevention strategies.

While certain pre-existing medical conditions and unhealthy lifestyle patterns are linked to more severe COVID-19 infection [7,8], disparities in COVID-19 outcomes have also been driven by numerous sociodemographic factors including age, sex, race, ethnicity, current socioeconomic status, and occupation [7,9,10]. For example, compared to White individuals, COVID-19 hospitalization in England is four times higher for Black individuals and two times higher for Asian individuals [9]. Similarly increased risk has been noted for people from lower socioeconomic status backgrounds, with these effects persisting even after accounting for lifestyle risk factors and other potential confounders [10]. Interestingly, nearly all of this research has examined relations between COVID-19 outcomes and contemporaneous sociodemographic variables, failing to consider early sociodemographic factors and neglecting developmental perspectives on the origins of health and disease [11]. To our knowledge, no studies have examined if COVID-19 outcomes are influenced by exposure to childhood adversity.

This knowledge gap is significant given that large-scale, epidemiological studies indicate that childhood adversity is associated with lifelong physical health disparities and early mortality [12,13]. For example, in a meta-analysis of 253,719 participants, those with high levels of childhood adversity were two-to-three times more likely to be diagnosed with heavy alcohol use, cancer, heart disease, and respiratory disease, compared to those with low levels of childhood adversity [13]. Similarly, a population-based cohort study of over one million individuals between 16 and 34 years of age found that those with the highest levels of childhood adversity had an all-cause mortality risk 4.5 times higher than those with no adversity; this mortality risk corresponded to 10.3 additional deaths (per 10,000 person-years) [14]. This increased mortality and morbidity risk is perhaps not surprising, given that childhood adversity relates to higher levels of inflammation and dysregulation of the hypothalamic pituitary adrenal axis [15,16]. With several studies now connecting excessive inflammation to COVID-19 disease severity and death [17], childhood adversity could be related to heightened negative outcomes related to COVID-19 through proinflammatory pathways or potentially other indirect mechanisms (i.e., unhealthy habits later in life [18]).

The current study seeks to investigate the association between early childhood adversity and COVID-19 mortality and morbidity in the UK Biobank, a large-scale and well characterized cohort. With childhood adversity being commonly linked to excessive inflammation and greater prevalence of negative health outcomes, we predicted that higher levels of adversity would be associated with higher rates of COVID-19-related hospitalization. Additionally, given work finding childhood adversity is related to early mortality overall, we hypothesized that adversity would be related to higher rates of COVID-19-related mortality. Finally, we anticipated that adjusting for potential confounds would reduce the strength of relations, but that childhood adversity would still be significantly associated with COVID-19-related negative outcomes.

## METHODS

### Study Design and Participants

The UK Biobank (UKBB) is a large-scale, biomedical research project focused on identifying risk factors for common life-threatening and disabling conditions in middle- and older-aged individuals. The UKBB database contains in-depth demographic, behavioral, and medical data from over half a million volunteer participants in the United Kingdom. At its onset in 2006, UKBB recruited residents between the ages of 40 and 69 that were registered with the United Kingdom’s National Health Service (NHS) and lived within 25 miles of an assessment site. Across a total of 22 assessment sites, participants completed touch-screen questionnaires and face-to-face interviews to collect information about their demographic backgrounds and lifestyles, including their ethnicity, level of education, weight and height measurements, chronic health conditions, and other variables [10,19]. Recruitment was completed in 2010, along with consent for future contact and linkage to routinely collected health-related data, such as those produced by the NHS. All UK Biobank participants provided informed consent electronically and the study was approved by the Northwest Multi-center Research Ethics Committee. Demographics for participants with different relevant data are shown in Table 1. Further details on data linkages, cleaning, validation and data availability (including summary statistics for all data fields) can be found on the UK Biobank data showcase webpage (https://biobank.ctsu.ox.ac.uk/crystal/).

**Table 01.**
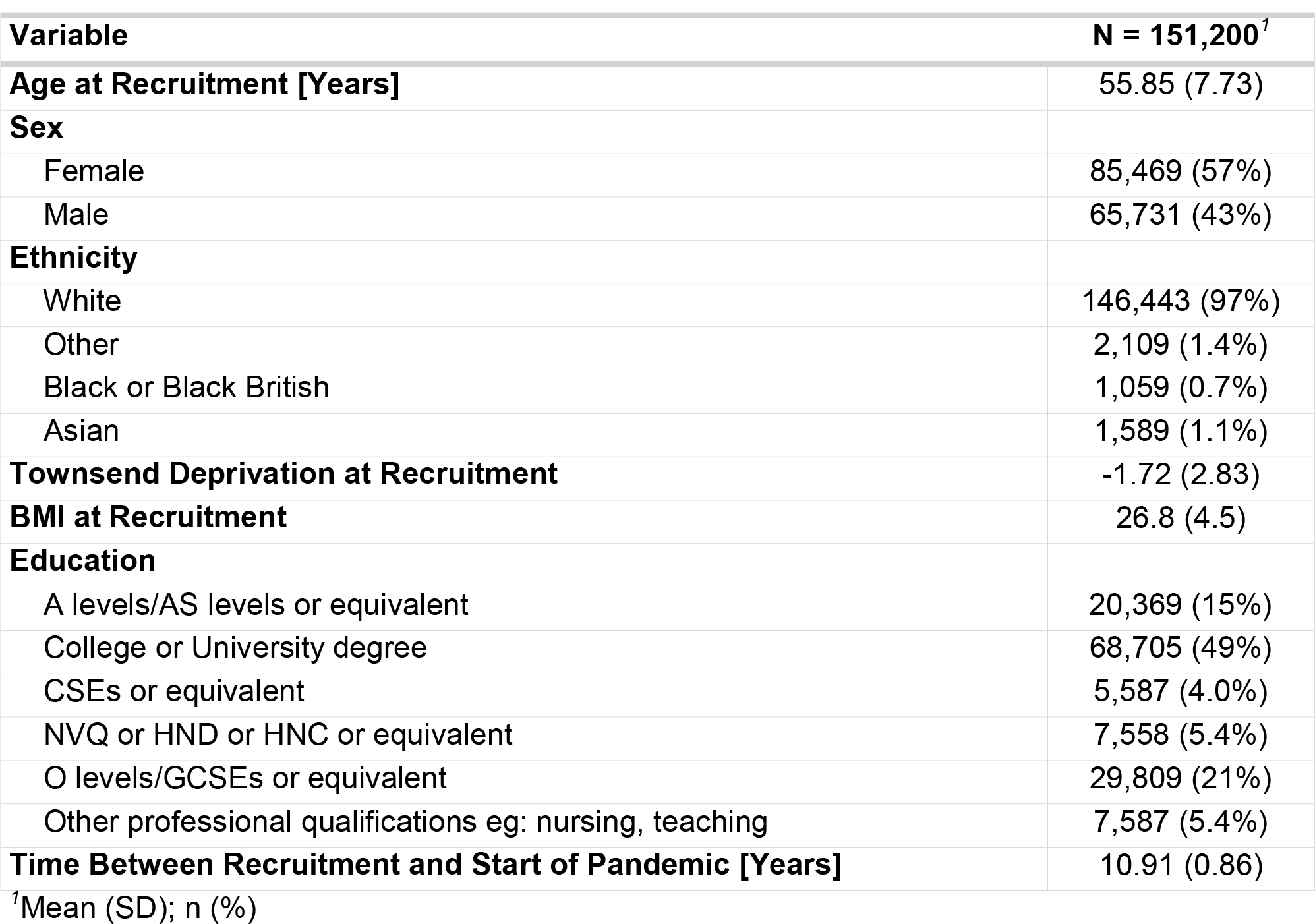
Demographic Table of Participants with usable data, listing age at recruitment, sex, ethnicity, socioeconomic status [Townsend Deprivation Index], body mass index [BMI], education, and time between recruitment and the start of the COVID-19 pandemic. Of note, all covariates (i.e., age, BMI) were measured at baseline when individuals began participation in the UKBB.

**Table 02.** The multivariate output for models where COVID-19-related hospitalization was the dependent variable (both panels) and sex, age, and ethnicity were the independent variables (left pane) or sex, age, ethnicity, and childhood adversity were the independent variables (right panel).

| *Variable* | **Base Model [without Adversity]:**<br>**Predicting Hospitalization** |  |  | **Model without Adversity**<br>**Predicting Hospitalization** |  |  |
| --- | --- | --- | --- | --- | --- | --- |
|  | *Odds Ratios* | *Conf. Int (95%)* | *P-Value* | *Odds Ratios* | *Conf. Int (95%)* | *P-Value* |
| Intercept | 0.00 | 0.00 – 0.00 | **<0.001** | 0.00 | 0.00 – 0.00 | **<0.001** |
| Ethnicity: Other [White Ref.] | 2.54 | 1.64 – 3.95 | **<0.001** | 2.28 | 1.47 – 3.55 | **<0.001** |
| Ethnicity: Black [White Ref.] | 3.60 | 2.09 – 6.20 | **<0.001** | 3.06 | 1.77 – 5.29 | **<0.001** |
| Ethnicity: Asian [White Ref.] | 2.13 | 1.24 – 3.64 | **0.006** | 1.93 | 1.13 – 3.31 | **0.017** |
| Sex [Female Reference] | 2.10 | 1.80 – 2.45 | **<0.001** | 2.15 | 1.84 – 2.51 | **<0.001** |
| Age (at Recruitment) | 1.26 | 1.17 – 1.36 | **<0.001** | 1.28 | 1.18 – 1.38 | **<0.001** |
| Childhood Adversity |  |  |  | 1.23 | 1.15 – 1.31 | **<0.001** |
| **Random Effects** |  |  |  |  |  |  |
| $\sigma^2$ | 3.29 | | | 3.29 | | |
| $\tau_{00}$ | 0.10<sub>site</sub> | | | 0.10<sub>site</sub> | | |
| ICC | 0.03 |  |  | 0.03 |  |  |
| N | 22<sub>site</sub> |  |  | 22<sub>site</sub> |  |  |
| Observations | 151200 |  |  | 151200 |  |  |
| Marginal R<sup>2</sup> / Conditional R<sup>2</sup> | 0.061 / 0.090 |  |  | 0.073 / 0.100 |  |  |

**Table 03.**
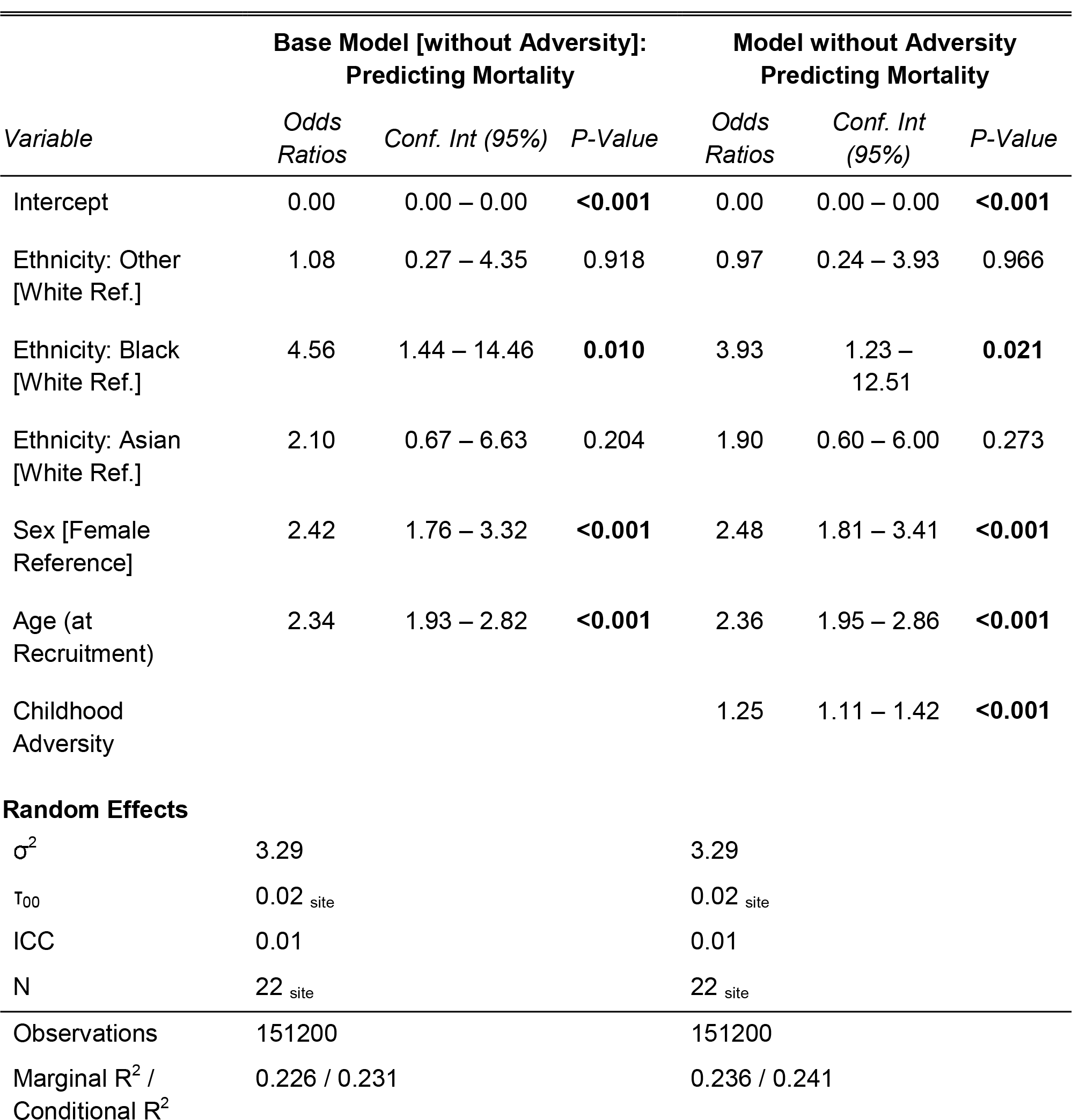
The multivariate output for models where COVID-19-related mortality was the dependent variable (both panels) and sex, age, and ethnicity were the independent variables (left pane) or sex, age, ethnicity, and childhood adversity were the independent variables (right panel).

For this study, we focused on four major data sources: 1) self-reports of childhood adversity; 2) COVID-19 outcomes, specifically hospitalization or death connected to COVID-19 infection; 3) demographic covariates (e.g., age, ethnicity); and 4) additional health-related variables (e.g., Body Mass Index) for use in sensitivity modeling. We detail information about each of these data sources below. Limiting participants to those with usable data in these categories, the average age in our sample was 55.91 years (+/- standard deviation=7.73), 43.68% male, and majority White (97%). Related to migration, the vast majority participants self-reported being born in the United Kingdom (93.18%). Additional demographics are listed in Table 1. A flow diagram depicting the number of participants included in our analyses, as well as information about participant exclusion, are shown in Figure 1.

**Figure 01.**
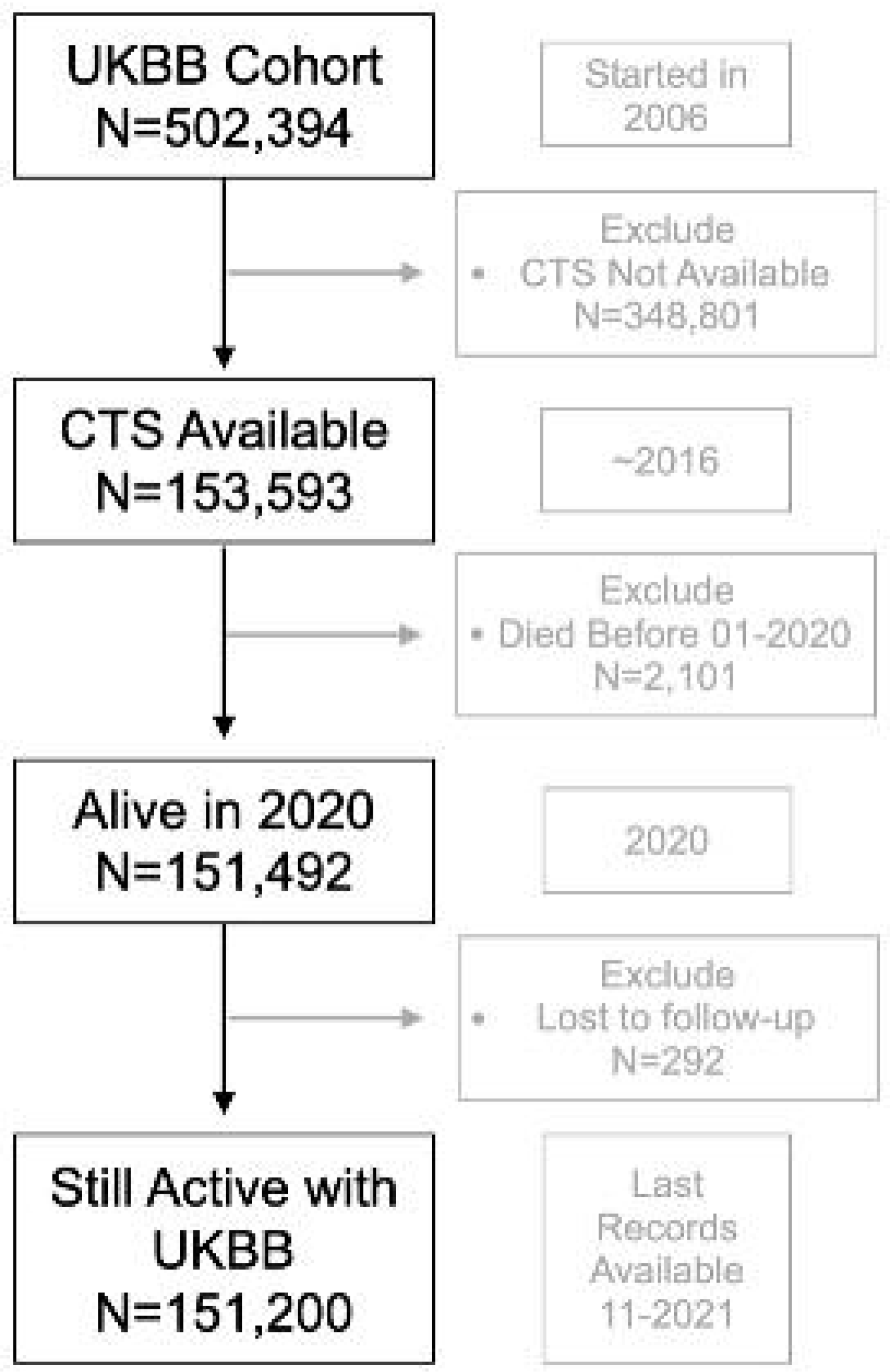
Flow diagram showing participants that were included (or excluded) from analyses based on data availability. The largest source of data loss was lack of childhood adversity variables (the Childhood Trauma Screen, CTS).

#### Research Ethics Approval

This work was approved by the University of Pittsburgh’s IRB (under protocol STUDY22080014).

### Self-Reports of Childhood Adversity

UKBB participants completed the Childhood Trauma Screener (CTS) [20], a shortened version of the Childhood Trauma Questionnaire, in an online follow-up after initial recruitment. The CTS is a 5-item questionnaire that asks about multiple forms of child maltreatment including physical abuse, physical neglect, emotional abuse, emotional neglect, and sexual abuse. Participants rated the frequency with which they felt loved or hated, were physically abused or sexually molested, and if someone took them to a doctor when they were children. Responses were made on a 5-point Likert scale from 0 [“*Never True*”] to 4 [“*Very Often True*”]. Participants could also select “*Prefer Not to Answer*” for any of the items. The CTS has been validated against other retrospective measures of child maltreatment and shows strong criterion and convergent validity, as well as good internal consistency [20]. The exact items of the CTS are noted in our supplemental materials. Of note, the original UKBB sample is 502,394 participants, but only 151,200 individuals had valid data for this questionnaire, were alive at the start of the COVID pandemic, and still active in the UKBB. As noted in our supplemental materials, participants in our analytic sample tended to be younger and more affluent (as indexed by lower Townsend Deprivation Index Scores). Our analytic subsample also was less diverse (including more White participants), more educated, and included more females than the full UKBB cohort. Full details about these analyses are included in our supplemental materials.

### COVID-19-Related Health Outcomes

We examined two classes of health outcomes related to COVID-19: 1) death where COVID-19 was reported as a primary or contributory cause and 2) inpatient hospitalization where NES data indicated COVID-19 occurrence. Both of these outcomes are related to International Classification of Diseases (ICD) code of U07.1: Confirmed COVID case [21]. Deaths were recorded through linkage to national death registries (NHS Digital, NHS Central Register, National Records of Scotland).

Of the 502,394 volunteers enrolled in the UK Biobank study, 69,444 died before 2020-01-10, the start of the COVID-19 pandemic as demarcated by the World Health Organization. These participants were therefore excluded from all analyses. Among the 432,950 participants alive at the start of the pandemic, 151,200 had previously completed the CTS and were the focus of analysis. Filtering for any occurrence of “*COVID*” in causes of death, 176 participants with complete childhood adversity data died due to COVID-19. In regard to hospitalization, 693 participants with complete childhood adversity data had an inpatient hospitalization related to COVID-19 (ICD Code U07.1). Health records were available up until November of 2021.

### Demographic Covariates

Different potential confounding factors were included in our statistical models. Initially, these included sex, ethnicity, and age at recruitment. Sex was classified as male or female; categories for ethnicity were White, Black or Black British, Asian or Asian British, Multi-racial, and Other ethnic group. Three additional sociodemographic and physical health factors were also examined: 1) Townsend Deprivation Index, an aggregated measure of socioeconomic status that quantifies the poverty level of an individual’s neighborhood using data on unemployment, car and home ownership, and household overcrowding that are associated with a particular postal code [22,23]; 2) Body Mass Index (BMI), derived from participants’ weight and height measurements; and 3) Chronic health conditions, a binary count of 10 self-reported diseases or serious medical issues. These included high blood pressure, diabetes, angina, hay fever, rhinitis or eczema, asthma, heart attack, emphysema/chronic bronchitis, deep-vein thrombosis (DVT, blood clot in leg), and stroke. Because the timing varied in regard to when participants were recontacted by UKBB and when information about potential confounding factors was available, we used sociodemographic and health data collected at recruitment. This was to minimize missing data, as only a portion of the sample would have updated data available (i.e., only deceased, or hospitalized participants would have updated age-data).

### Statistical Modeling

To understand the impact of childhood adversity on COVID-19 outcomes, we used mixed effects logistic regression analysis to generate odds ratios (ORs) with 95% confidence intervals (CI). As noted previously, our final analytic sample was 151,200 participants. These individuals had valid measurements of childhood adversity, were alive at the start of the COVID pandemic, and still active in the UKBB. With each outcome (COVID-19-related hospitalization; COVID-19-related mortality), four model variations were used: a) sex, age, and ethnicity were included as independent variables, without inclusion of childhood adversity (model 1); b) sex, age, ethnicity, and childhood adversity as independent variables (model 2); c) sex, age, ethnicity, current socioeconomic status [Townsend Deprivation Index] and childhood adversity as independent variables (model 3); and d) sex, age, ethnicity, current socioeconomic status, physical health history [Chronic health conditions] and childhood adversity as independent variables (model 4). This allowed us to first understand risks caused by childhood adversity (model 2), while iteratively eliminating the effects of potential confounders and/or mediators (models 3 and 4) to understand the role of demographic and physical comorbidities in attenuating risks. The outcome variable in each model (COVID-19-related hospitalization, or COVID-19-related mortality) was a binary indicator of the occurrence of that negative health outcome (1=COVID-19 related hospitalization, or COVID-19-related mortality; 0=no hospitalization, or mortality). Independent variables were deemed significant based on their *p*-values, as well as 95% confidence intervals of ORs that crossed 1. When appropriate, we also compared different models using Binomial ANOVAs with *p*-values calculated using a *χ*^2^ test and *χ*^2^ distribution.

For statistical modeling, we used R version 4.2.2 and the following R packages: aod v. 1.3.2, broom.mixed v. 0.2.9.4, car v. 3.1.1, cowplot v. 1.1.1, gt v. 0.8.0, gtsummary v. 1.6.3, hrbrthemes v. 0.8.0, jtools v. 2.2.1, lme4 v. 1.1.31, modelbased v. 0.8.6, performance v. 0.10.1, plyr v. 1.8.8, and tidyverse v. 1.3.2.

## RESULTS

Across multiple statistical models, we observed an association between childhood adversity and COVID-19 related hospitalization. For all models, higher self-reports of childhood adversity were related to greater likelihood of COVID-19-related hospitalization. In our model adjusted for age, ethnicity, and sex (model 2), childhood adversity was associated with an OR of 1.227 of hospitalization (95% CI=1.153-1.306, Childhood Adversity *z*=6.49, *p*<0.005). Notably, compared to our model that only included age, ethnicity, and sex (model 1), inclusion of childhood adversity improved model fit (*χ*^2^ (1, N=151200)=36.5, *p*<0.005) with an improvement in R^2^ (Model 1 Conditional R^2^: 0.090, AIC= 8669.9; Model 2 Conditional R^2^: 0.100, AIC= 8635.4). Receiver operating characteristic curves and regression coefficient plots for these models are shown in Figure 02.

**Figure 02.**
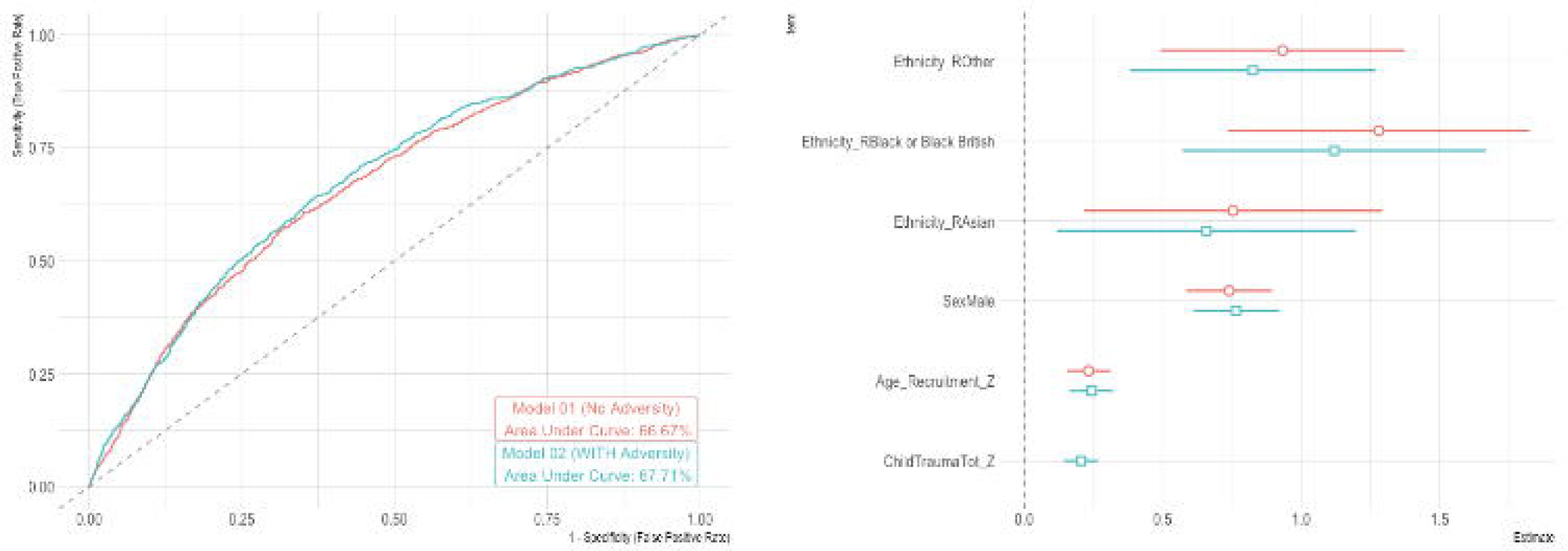
This two-panel figure shows results for COVID-19 hospitalization. Panel A shows a receiver operating characteristic curve for two models [Model 01: Age, sex, and ethnicity predicting hospitalization, but no measure of childhood adversity included in our mixed effects logistic regression analysis; Model 2: Age, sex, ethnicity, and childhood adversity predicting hospitalization]. Panel B shows regression coefficients from these mixed effects logistic regression analysis, with sex, age, and childhood adversity depicted. Ethnicity was omitted. Model 01 is colored orange, while Model 02 is colored teal.

Adjustment for potential confounds attenuated this association, but still suggested a connection between childhood adversity and COVID-19-related hospitalization. In models adjusting for age, ethnicity, sex, and current socioeconomic status (model 3), childhood adversity was associated with an OR of 1.20 of hospitalization (95% CI=1.13-1.28, Childhood Adversity *z*=5.80, *p*<0.005). When we also include chronic health conditions (model 4), the association between childhood adversity and COVID-19 related hospitalization remains mostly unchanged (OR=1.193, 95% CI=1.121-1.269, Childhood Adversity *z*=5.55, *p*<0.005).

Related to COVID-19-related mortality, childhood adversity was associated with an OR of 1.25 of a COVID-19 related death (95% CI=1.11-1.424, Childhood Adversity *z*=3.50, *p*<0.005) in our models adjusting for age, ethnicity, and sex (model 2). Notably, compared to our model that only included age, ethnicity, and sex (model 1), inclusion of childhood adversity improved model fit (*χ*^2^ (1)=10.5, *p*=0.001) with an improvement in R^2^ (Model 1 Conditional R^2^ =0.231 AIC= 2606.0; Model 2 Conditional R^2^= 0.241, AIC=2597.5). Receiver operating characteristic curves and regression coefficient plots for these models are shown in Figure 03.

**Figure 03.**
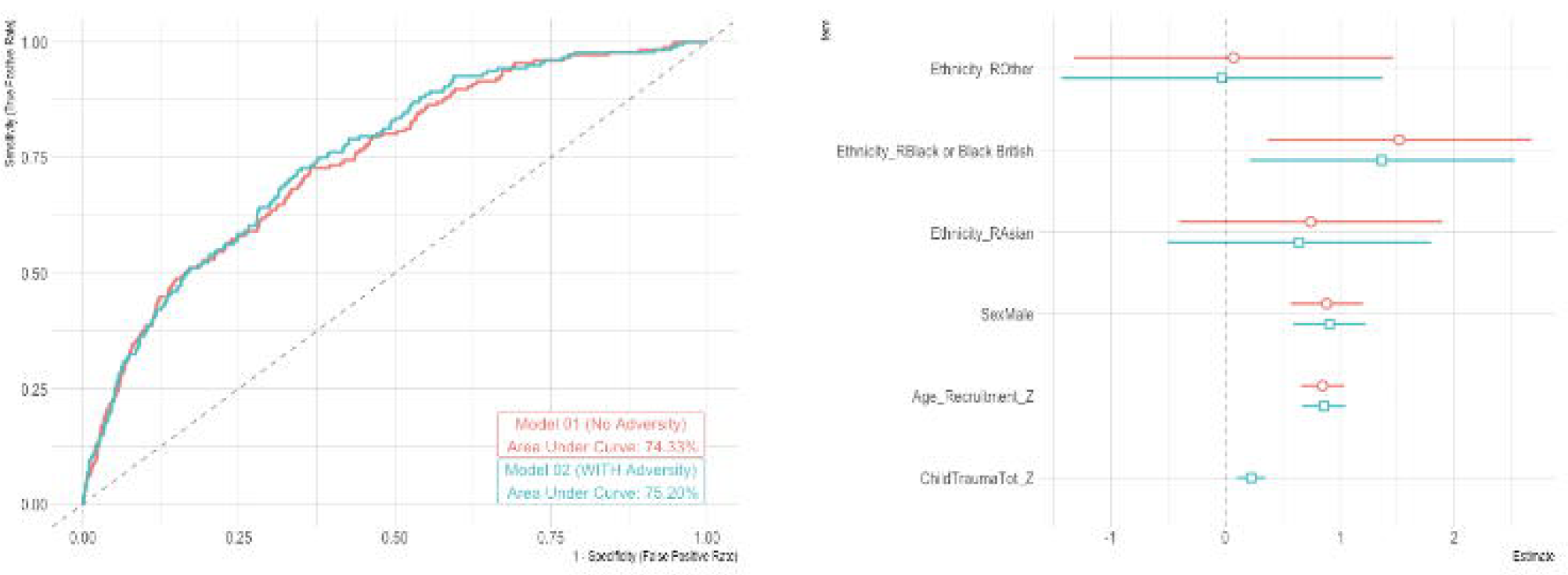
This two-panel figure shows results for COVID-19 mortality. Panel A shows a receiver operating characteristic curve for two models [Model 01: Age, sex, and ethnicity predicting mortality, but no measure of childhood adversity included in our mixed effects logistic regression analysis; Model 2: Age, sex, ethnicity, and childhood adversity predicting mortality]. Panel B shows regression coefficients from these mixed effects logistic regression analysis, with sex, age, and childhood adversity depicted. Ethnicity was omitted. Model 01 is colored orange, while Model 02 is colored teal.

Again, confound adjustment attenuated associations, but statistical models still suggested a connection between childhood adversity and COVID-19 related mortality. In models adjusting for age, ethnicity, sex, and current socioeconomic status (model 3), childhood adversity was associated with an OR of 1.214 of hospitalization (95% CI=1.069-1.379, Childhood Adversity *z*=2.99, *p*=0.003). When we also include chronic health conditions (model 4), the association between childhood adversity and COVID-related mortality remains mostly unchanged (OR=1.204, 95% CI=1.061-1.367, Childhood Adversity *z*=2.88, *p*=0.004). Additional sensitivity models examining additional confounding factors (i.e., migration status) and interactions between chronic conditions and childhood adversity are noted in our Supplemental Materials. In our Supplement, we also complete exploratory analyses examining the potential mechanisms linking childhood adversity to COVID outcomes. This involved indirect (“mediation”) models where we tested whether statistical associations between adversity (X) and COVID mortality or hospitalization outcomes (Y) was reduced when accounting for current socioeconomic status or pre-existing health conditions (M).

## DISCUSSION

In a large-scale, well-characterized cohort, we found links between childhood adversity and COVID-19-related outcomes. Specifically, we found significant associations between childhood adversity and both COVID-19-related hospitalization and COVID-19-related mortality. For both morbidity and mortality, these links were seen in statistical models adjusted for important sociodemographic and physical health confounders, underscoring the significance of childhood adversity in predicting mortality and morbidity risk.

Our data fills in important knowledge gaps both in terms of childhood adversity and health, as well as risk factors related to COVID-19-related hospitalization and mortality. The results detailed here align with past work linking higher childhood adversity to poor physical health, including heavy alcohol use, cancer, heart disease, and respiratory disease. Similarly, previous studies have found that particularly high levels of adversity are connected to higher all-cause mortality risk. Related to these two bodies of work, multiple meta-analyses suggest childhood adversity connects to higher levels of inflammation. While this investigation was unable to speak to potential mechanisms, it is likely that higher levels of inflammation, as well as alterations in the hypothalamic pituitary adrenal axis, related to childhood adversity are connected to the increased mortality and hospitalization observed here. Alternatively, childhood adversity may exacerbate the multitude of stressors associated with the pandemic, such as social isolation, economic hardship, and health concerns, further increasing COVID-19-related mortality and morbidity.

While this is the first study to examine links between childhood adversity and COVID-19-related outcomes in a large cohort, this project is not without limitations. First, our measure of childhood adversity was only available on a modest proportion of the overall UKBB sample (∼30.1%). This may skew results as perhaps only certain individuals completed this measure. In addition, the UKBB is not truly a nationally representative cohort, with a response rate of ∼5.5%. Participants tend to live in less socioeconomically deprived areas and are predominantly Caucasian. Associations between childhood adversity and COVID-19-related outcomes could differ in socioeconomically deprived areas, as well as in communities of color. Second, while we controlled for many important, potential confounds, these were measured at the baseline assessment of the UKBB cohort. Baseline assessments were conducted multiple years before the COVID-19 pandemic and participants’ current health or lifestyle may differ from when they started in the study. Notably, previous studies suggest UKBB baseline data can accurately rank participants years later [24], but this has only been investigated for a few outcomes in the project. Third, the examination of morbidity and mortality may be an imperfect assessment of COVID-19-related outcomes. Hospitalization data, for example, listed COVID-19 as a primary reason for admittance, but there may have been other diseases driving hospitalization (e.g., pneumonia). There may also be misclassification of deaths and hospitalizations due to COVID; future work could potentially probe multiple indices of health to understand if COVID is the true cause of hospitalization or mortality. Of note, COVID-19 infection was not necessarily confirmed in each case. Furthermore, data was only present until the end of 2021; different strains of COVID-19 have surged around the globe and may be associated with differential mortality and morbidity, a possibility these data are unable to address. Lastly, in considering our statistical models, links between adversity and COVID-19-related mortality were modest in magnitude, though still statistically significant.

These results also further delineate the sociodemographic and psychological factors contributing to COVID-19-related negative outcomes. Clear from past work is that certain pre-existing medical conditions and unhealthy lifestyle patterns are linked to more severe COVID-19 infection, which subsequently contributes to an increased likelihood of hospitalization and mortality. Notably, our work extends past studies that have shown that sociodemographic risk factors are significant drivers of COVID-19 disparities. While previous projects have found that age, sex, race and ethnicity, and current socioeconomic status increase negative outcomes related to COVID-19, we believe this is the first project to examine how childhood adversity may further amplify risk. While the medical and public health communities have raised awareness about how sociodemographic variables may influence the impact of COVID-19, our work underscores that it is also critical to consider how an individual’s developmental history may heighten the impact of the pandemic.

The association between experiences of childhood adversity and COVID-19 morbidity and mortality emphasize the need for further work considering modifiable and more proximal psychosocial factors. Future work could investigate if psychological processes related to adversity, such as depression, self-concept, or self-regulation, cascade from childhood experiences to the adult health outcomes that we investigated here. Pinpointing these processes may allow for policy and interventions to lessen the negative impact of COVID-19 in those that have suffered childhood adversity. Further work in this space will be critical to reduce adversity-related negative outcomes with COVID-19, particularly as this disease becomes endemic, and to limit adversity-related negative outcomes with future pandemics.

## Supporting information

Supplemental Materials/Analyses

## Data Availability

All data are available from the UB Biobank. This resource is available to scientists from the UK and outside, whether they work in the public or private sector, for industry, academia or a charity, subject to verification that the research is health-related and in the public interest.

## REFERENCES

1 World Health Organization. WHO Coronavirus (COVID-19) Dashboard. 2023.https://covid19.who.int (accessed 24 Jan 2023).

2 Schöley J, Aburto JM, Kashnitsky I, et al. Life expectancy changes since COVID-19. Nat Hum Behav 2022;6:1649–59. doi:10.1038/s41562-022-01450-3

3 Wang H, Paulson KR, Pease SA, et al. Estimating excess mortality due to the COVID-19 pandemic: a systematic analysis of COVID-19-related mortality, 2020–21. The Lancet 2022;399:1513–36.

4 Bartsch SM, Ferguson MC, McKinnell JA, et al. The Potential Health Care Costs And Resource Use Associated With COVID-19 In The United States: A simulation estimate of the direct medical costs and health care resource use associated with COVID-19 infections in the United States. Health affairs 2020;39:927–35.

5 Douaud G, Lee S, Alfaro-Almagro F, et al. SARS-CoV-2 is associated with changes in brain structure in UK Biobank. Nature 2022;604:697–707. doi:10.1038/s41586-022-04569-5

6 Xie Y, Xu E, Bowe B, et al. Long-term cardiovascular outcomes of COVID-19. Nat Med 2022;28:583–90. doi:10.1038/s41591-022-01689-3

7 Elliott J, Bodinier B, Whitaker M, et al. COVID-19 mortality in the UK Biobank cohort: revisiting and evaluating risk factors. European journal of epidemiology 2021;36:299–309.

8 Ko JY, Danielson ML, Town M, et al. Risk Factors for Coronavirus Disease 2019 (COVID-19)–Associated Hospitalization: COVID-19–Associated Hospitalization Surveillance Network and Behavioral Risk Factor Surveillance System. Clinical Infectious Diseases 2021;72:e695–703. doi:10.1093/cid/ciaa1419

9 Lassale C, Gaye B, Hamer M, et al. Ethnic disparities in hospitalisation for COVID-19 in England: The role of socioeconomic factors, mental health, and inflammatory and pro-inflammatory factors in a community-based cohort study. Brain, Behavior, and Immunity 2020;88:44–9. doi:10.1016/j.bbi.2020.05.074

10 Foster HM, Ho FK, Mair FS, et al. The association between a lifestyle score, socioeconomic status, and COVID-19 outcomes within the UK Biobank cohort. BMC infectious diseases 2022;22:1–13.

11 Gluckman PD, Hanson MA, Cooper C, et al. Effect of in utero and early-life conditions on adult health and disease. New England journal of medicine 2008;359:61–73.

12 Elsenburg LK, Rieckmann A, Nguyen T-L, et al. Mediation of the parental education gradient in early adult mortality by childhood adversity: a population-based cohort study of more than 1 million children. The Lancet Public Health 2022;7:e146–55.

13 Hughes K, Bellis MA, Hardcastle KA, et al. The effect of multiple adverse childhood experiences on health: a systematic review and meta-analysis. The Lancet Public Health 2017;2:e356–66.

14 Rod NH, Bengtsson J, Budtz-Jørgensen E, et al. Trajectories of childhood adversity and mortality in early adulthood: a population-based cohort study. The Lancet 2020;396:489–97.

15 Chiang JJ, Lam PH, Chen E, et al. Psychological stress during childhood and adolescence and its association with inflammation across the lifespan: A critical review and meta-analysis. Psychological Bulletin 2022;148:27.

16 Kuhlman KR, Horn SR, Chiang JJ, et al. Early life adversity exposure and circulating markers of inflammation in children and adolescents: A systematic review and meta-analysis. Brain, behavior, and immunity 2020;86:30–42.

17 Merad M, Martin JC. Pathological inflammation in patients with COVID-19: a key role for monocytes and macrophages. Nature reviews immunology 2020;20:355–62.

18 Brugiavini A, Buia R, Kovacic M, et al. Adverse childhood experiences and unhealthy lifestyles later in life: evidence from SHARE countries. Review of Economics of the Household 2022;:1–18.

19 Jani BD, Hanlon P, Nicholl BI, et al. Relationship between multimorbidity, demographic factors and mortality: findings from the UK Biobank cohort. BMC medicine 2019;17:1–13.

20 Grabe HJ, Schulz A, Schmidt CO, et al. A brief instrument for the assessment of childhood abuse and neglect: the childhood trauma screener (CTS). Psychiatrische Praxis 2012;39:109–15.

21 World Health Organization. Emergency use ICD codes for COVID-19 disease outbreak. https://www.who.int/standards/classifications/classification-of-diseases/emergency-use-icd-codes-for-covid-19-disease-outbreak (accessed 14 Feb 2023).

22 Phillimore P, Beattie A, Townsend P. Health and deprivation. inequality and the North. London: Croom Helm. Health Policy 1988;10:207–206.

23 Townsend P. Deprivation. Journal of Social Policy 1987;16:125–46. doi:10.1017/S0047279400020341

24 Bradbury KE, Young HJ, Guo W, et al. Dietary assessment in UK Biobank: an evaluation of the performance of the touchscreen dietary questionnaire. Journal of nutritional science 2018;7:e6.

