## Supplemental Materials/Analyses for "Childhood Adversity and COVID-19 Outcomes in the UK Biobank"

***Full Multivariate Model Outputs***

Connected to models in the main manuscript, Table S1 shows multivariate output for models where COVID-19-related hospitalization was the dependent variable, and sex, age, chronic health conditions, socioeconomic deprivation, and ethnicity were the independent variables or sex, age, chronic health conditions, socioeconomic deprivation, ethnicity and childhood adversity were the independent variables. Table S2 shows multivariate output for models where COVID-19-related mortality was the dependent variable, and sex, age, chronic health conditions, socioeconomic deprivation, and ethnicity were the independent variables or sex, age, chronic health conditions, socioeconomic deprivation, ethnicity and childhood adversity were the independent variables.

***Specific Questions Related to Childhood Adversity***

Our approach is anchored in the broad tradition of the “Adverse Childhood Experiences” (ACEs) studies. Initially, that study examined childhood trauma and later-life health outcomes in over 17,000 participants from the state of California in the United States. The ACEs study found a strong correlation between adverse childhood experiences like abuse, neglect, and household dysfunction, and poor health and quality of life outcomes in adulthood. Here to assess childhood adversity, we used the Childhood Trauma Screener (CTS), a 5-item questionnaire that asks about multiple forms of child maltreatment including physical abuse, physical neglect, emotional abuse, emotional neglect, and sexual abuse. In this sample, participants were asked “When I was growing up:”

- I Felt loved as a child
- People in my family hit me so hard that it left me with bruises or marks
- I felt that someone in my family hated me
- Someone molested me (sexually)
- There was someone to take me to the doctor if I needed it

***Sample Characteristics of Individuals with and without Childhood Adversity Measures***

As noted in our main manuscript, the Childhood Trauma Screener (CTS) was only available in ~30% of the UK Biobank cohort. This could be creating biases in those who completed this questionnaire compared to those who refused to complete the CTS. To investigate potential biases, we constructed multiple statistical models to examine potential differences; for continuous variables (i.e., age at recruitment; Townsend Deprivation Index), we used generalized linear mixed models for binomial data (CTS availability coded as 0=not available; 1=available). These models included a random factor for site and each model only included one independent variable (i.e., age at recruitment; Townsend Deprivation Index). For sociodemographic variables that were dichotomous or categorical (i.e., sex; education), we used chi-square test of independence, comparing the observed frequencies of each combination of the categories for CTS availability (again, CTS availability coded as 0=not available; 1=available). Of note, these models did not include a variable for site and each model only included one categorical variable (i.e., sex; education),

These models revealed that participants in our analytic sample (those with valid data for this questionnaire, were alive at the start of the COVID pandemic, and still active in the UKBB) were significantly younger (Age at Recruitment, z=20.51, p<.005) and more affluent (as indexed by Townsend Deprivation Index Scores z=68.23, p<.005). These effects are shown in Figure S1. Analysis of categorical variables indicated that the analytic sample (again, who completed the CTS) compared to the excluded participants (who did not completed the CTS) included more females (Sex χ²(1)=221.46, p<.005), was less diverse (including more White participants; Race/Ethnicity χ²(5)=3465, p<.005), more educated (Education χ²(5)=3465, p<.005), and contained fewer individuals that migrated to the United Kingdom (Migration χ²(1)=835.12, p<.005). These effects are shown in Figures S2-S3. Clear from these results is that there may be biases operating in our analyses. We would, however, argue that such limitations are causing a potential underestimation of effects. All of these factors are likely to increase COVID mortality and morbidity, but our cohort has lower levels of risk (i.e., more affluence; education, etc.). As such, the true relations between childhood adversity and these outcomes could be stronger than we report in the main manuscript.

***Exploratory Mediation Models Examining Potential Important Indirect Pathways from Adversity to COVID Outcomes***

To understand the potential mechanisms linking childhood adversity to COVID outcomes, we utilized indirect (“mediation”) models. These models examined if statistical associations between adversity (X) and COVID mortality or hospitalization outcomes (Y) was reduced when accounting for current socioeconomic status or pre-existing health conditions (M). This was done by examining if there was a relation between adversity and current socioeconomic status (Townsend Deprivation Index Scores, a1 path) or pre-existing health conditions (a binary count of 10 self-reported diseases or serious medical issues, a2 path); then we probed if there were relations between current socioeconomic status (b1 path) and COVID mortality or hospitalization, as well as pre-existing health conditions (b2 path) and COVID mortality or hospitalization. Indirect effects, as well as a 95% confidence interval, were then calculated (a1 xb1; a2 x b2) to test for “statistical mediation”. We used similar covariates to those in the main manuscript (e.g., age, sex, ethnicity [Person of color; not]) and also modeled the covariance between current socioeconomic status and pre-existing health conditions.

Related to COVID mortality, both current socioeconomic status and pre-existing health conditions indirectly explained links between adversity and this outcome. In these models, childhood adversity was related to current socioeconomic status (a1=0.110, z=46.511, p<0.005) and also pre-existing health conditions (a2=0.008, z=3.20, p=0.001). Current socioeconomic status and pre-existing health conditions were also both related to COVID mortality (current socioeconomic status (b1=0.076, z=3.938, p<0.005; and pre-existing health conditions b2=0.092, z=5.458, p<0.005). These indirect effects (a1 x b1, or a2 x b2) were both significant (current socioeconomic status indirect effect=0.008, z=3.925, p<0.005; pre-existing health conditions indirect effect=0.001, z=2.767, p=0.006). Of note, COVID mortality was related to childhood adversity (z=2.044, p=0.041). These effects are shown in Figure S3.

Connected to COVID hospitalization, both current socioeconomic status and pre-existing health conditions indirectly explained links between adversity and this outcome. Similar to the mortality models, childhood adversity was related to current socioeconomic status (a1=0.110, z=46.511, p<0.005) and also pre-existing health conditions (a2=0.008, z=3.20, p=0.001). Current socioeconomic status and pre-existing health conditions were also both related to COVID hospitalization (current socioeconomic status (b1=0.066, z=5.476, p<0.005; and pre-existing health conditions b2=0.082, z=8.411, p<0.005). These indirect effects (a1 x b1, or a2 x b2) were both significant (current socioeconomic status indirect effect=0.007, z=5.446, p<0.005; pre-existing health conditions indirect effect=3.006, z=2.767, p=0.003). Of note, COVID mortality was related to childhood adversity (z=5.331, p<0.005). These effects are shown in Figure S4.

Of note, we urge caution in interpreting these results. The UK Biobank measures childhood adversity retrospectively at (or near the same time) it asked participants about pre-existing health conditions. Since there is not temporal separation/independence of variables, this would actually run against fundamental assumptions of mediation (or “indirect effect”) models (as articulated by Ref. S1).

***Sensitivity Analyses Examining COVID Antibodies***

UK Biobank participants were invited to take part in a SARS-CoV-2 coronavirus antibody study during the introduction of the COVID vaccine in the United Kingdom. Specifically, participants were mailed lateral flow test kits to detect COVID antibodies. Those with positive results who were unvaccinated were sent a second kit to reduce potential false positives. Of note, these antibody tests couldn't distinguish between antibodies from infection vs vaccination. Participants with positive results were later invited to provide a blood sample to test specifically for IgM antibodies that are only produced after infection. Across the whole UK Biobank, ~200,000 participants took part in these follow-up procedures, but only 106,731 of these participants had valid measures of childhood adversity. In our analytic sample, 63997 participants tested negative for any COVID antibodies, 41962 tested positive for IgG antibodies (which could result from infection or vaccine), and 772 tested positive for IgM.

Equipped with this information, we then constructed mixed effects logistic regression analysis to generate odds ratios (ORs) with 95% confidence intervals (CI). Similar to our main manuscript (model 2), these models included sex, age, ethnicity, and childhood adversity as independent (fixed effect) variables, while site was included as a random effect. Because of timing variability in when antibody testing kits were sent and received, when vaccines were administered, and whether an individual was infected with COVID, we were unable to specifically isolate who had IgG anitbodies were due purely to infection (and not vaccination). This led us to examine COVID antibodies presence using three coding schemes: a) IgG and IgM were combined into a “Positive” group to maximize sample size; b) IgG was recoded as “Positive” group and IgM was omitted from analyses; and c) IgM was recoded as “Positive” group and IgG was omitted from analyses. In each of these models, individuals with negative antibody results were the reference group (coded as 0) and the different positive groups were coded as 1.

In our first model (where IgG and IgM antibodies were combined), childhood trauma was related to lower incidence of antibodies (OR=0.972, 95% CI of OR=0.959-0.984, Z=-4.401, p<.005, shown in Figure S5). In our second (where IgM values were omitted from analyses), childhood trauma was related to lower incidence of antibodies (OR=0.97, 95% CI=0.958-0.983, Z=-4.58, p<.005). In our second (where IgG values were omitted from analyses), childhood trauma was not related to incidence of antibodies (OR=1.04, 95% CI=0.967-1.12, Z=1.056, p=0.291). Of note, this had a much smaller sample (N=64,769, with only ~1% of participants in non-reference group [IgM antibody positive N=772). We would therefore urge caution in interpretation of effects given potential overdispersion in these logistic models.

***Analyses Examining Interactions Between Adversity and Chronic Health Issues***

To understand if pre-existing health issues and childhood adversity interactively drove COVID outcomes, we constructed mixed effects logistic regression analysis to generate OR with 95% CI where the interaction of these factors was modeled. This statistical approach was similar to model 4 in the main manuscript, but also included an additional independent (fixed effect). variable of pre-existing health issues X childhood adversity. We examined this in separate statistical models where COVID death or hospitalization was the dependent variable. For COVID death, this interaction term was not significant (OR=0.9657, 95% CI of OR=0.887-1.052, Z=-.80, p=0.423). Similarly, this interaction term was not significant in statistical models where COVID hospitalization was the dependent variable of interest (OR=0.984, 95% CI of OR=0.945-1.025, Z=-.78, p=0.434). of note, the main effects of pre-existing health issues and also childhood adversity remained significant in these models examining COVID death or hospitalization (all p’s<.005).

**References**

S1. Cole, D. A., & Maxwell, S. E. (2003). Testing mediational models with longitudinal data: questions and tips in the use of structural equation modeling. Journal of abnormal psychology, 112(4), 558.

Table S1.

|  | **Adjusted Model [without Adversity]: Predicting Hospitalization** | | | **Adjusted Model with Adversity Predicting Hospitalization** | | |
| --- | --- | --- | --- | --- | --- | --- |
| *Variable* | *Odds Ratios* | *Conf. Int (95%)* | *P-Value* | *Odds Ratios* | *Conf. Int (95%)* | *P-Value* |
| Intercept | 0.00 | 0.00 – 0.00 | **<0.001** | 0.00 | 0.00 – 0.00 | **<0.001** |
| Chronic Health Conditions | 1.25 | 1.18 – 1.32 | **<0.001** | 1.24 | 1.18 – 1.32 | **<0.001** |
| Socioeconomic Deprivation | 1.26 | 1.17 – 1.36 | **<0.001** | 1.23 | 1.14 – 1.32 | **<0.001** |
| Ethnicity: Other [White Ref.] | 2.36 | 1.51 – 3.67 | **<0.001** | 2.16 | 1.38 – 3.36 | **0.001** |
| Ethnicity: Black [White Ref.] | 2.96 | 1.71 – 5.12 | **<0.001** | 2.63 | 1.52 – 4.56 | **0.001** |
| Ethnicity: Asian [White Ref.] | 2.14 | 1.25 – 3.67 | **0.006** | 1.98 | 1.15 – 3.40 | **0.013** |
| Sex [Female Reference] | 2.08 | 1.78 – 2.43 | **<0.001** | 2.12 | 1.82 – 2.48 | **<0.001** |
| Age (at Recruitment) | 1.22 | 1.13 – 1.32 | **<0.001** | 1.23 | 1.14 – 1.34 | **<0.001** |
| Childhood Adversity |  |  |  | 1.19 | 1.12 – 1.27 | **<0.001** |
| **Random Effects** | | | | | | |
| σ^2^ | 3.29 | | | 3.29 | | |
| τ_00_ | 0.11 _site_ | | | 0.10 _site_ | | |
| N | 22 _site_ | | | 22 _site_ | | |
| Observations | 151006 | | | 151006 | | |
| Marginal R^2^ / Conditional R^2^ | 0.085 / 0.114 | | | 0.093 / 0.120 | | |

*Caption*: The multivariate output for models where COVID-19-related hospitalization was the dependent variable (both panels) and sex, age, chronic health conditions, socioeconomic deprivation, and ethnicity were the independent variables (left pane) or sex, age, chronic health conditions, socioeconomic deprivation, ethnicity and childhood adversity were the independent variables (right panel).

Table S2.

|  | **Adjusted Model [without Adversity]: Predicting Mortality** | | | **Adjusted Model with Adversity Predicting Mortality** | | |
| --- | --- | --- | --- | --- | --- | --- |
| *Variable* | *Odds Ratios* | *Conf. Int (95%)* | *P-Value* | *Odds Ratios* | *Conf. Int (95%)* | *P-Value* |
| Intercept | 0.00 | 0.00 – 0.00 | **<0.001** | 0.00 | 0.00 – 0.00 | **<0.001** |
| Chronic Health Conditions | 1.21 | 1.09 – 1.35 | **<0.001** | 1.20 | 1.08 – 1.34 | **0.001** |
| Socioeconomic Deprivation | 1.40 | 1.22 – 1.60 | **<0.001** | 1.37 | 1.20 – 1.57 | **<0.001** |
| Ethnicity: Other [White Ref.] | 0.93 | 0.23 – 3.75 | 0.914 | 0.85 | 0.21 – 3.47 | 0.826 |
| Ethnicity: Black [White Ref.] | 3.16 | 0.98 – 10.13 | 0.053 | 2.87 | 0.89 – 9.23 | 0.077 |
| Ethnicity: Asian [White Ref.] | 1.94 | 0.61 – 6.12 | 0.259 | 1.80 | 0.57 – 5.70 | 0.314 |
| Sex [Female Reference] | 2.40 | 1.75 – 3.30 | **<0.001** | 2.45 | 1.79 – 3.37 | **<0.001** |
| Age (at Recruitment) | 2.28 | 1.88 – 2.76 | **<0.001** | 2.30 | 1.90 – 2.79 | **<0.001** |
| Childhood Adversity |  |  |  | 1.20 | 1.06 – 1.37 | **0.004** |
| **Random Effects** | | | | | | |
| σ^2^ | 3.29 | | | 3.29 | | |
| τ_00_ | 0.01 _site_ | | | 0.01 _site_ | | |
| N | 22 _site_ | | | 22 _site_ | | |
| Observations | 151006 | | | 151006 | | |
| Marginal R^2^ / Conditional R^2^ | 0.245 / 0.247 | | | 0.252 / 0.254 | | |

Caption: The multivariate output for models where COVID-19-related mortality was the dependent variable (both panels) and sex, age, chronic health conditions, socioeconomic deprivation, and ethnicity were the independent variables (left pane) or sex, age, chronic health conditions, socioeconomic deprivation, ethnicity and childhood adversity were the independent variables (right panel).

Figure S1.

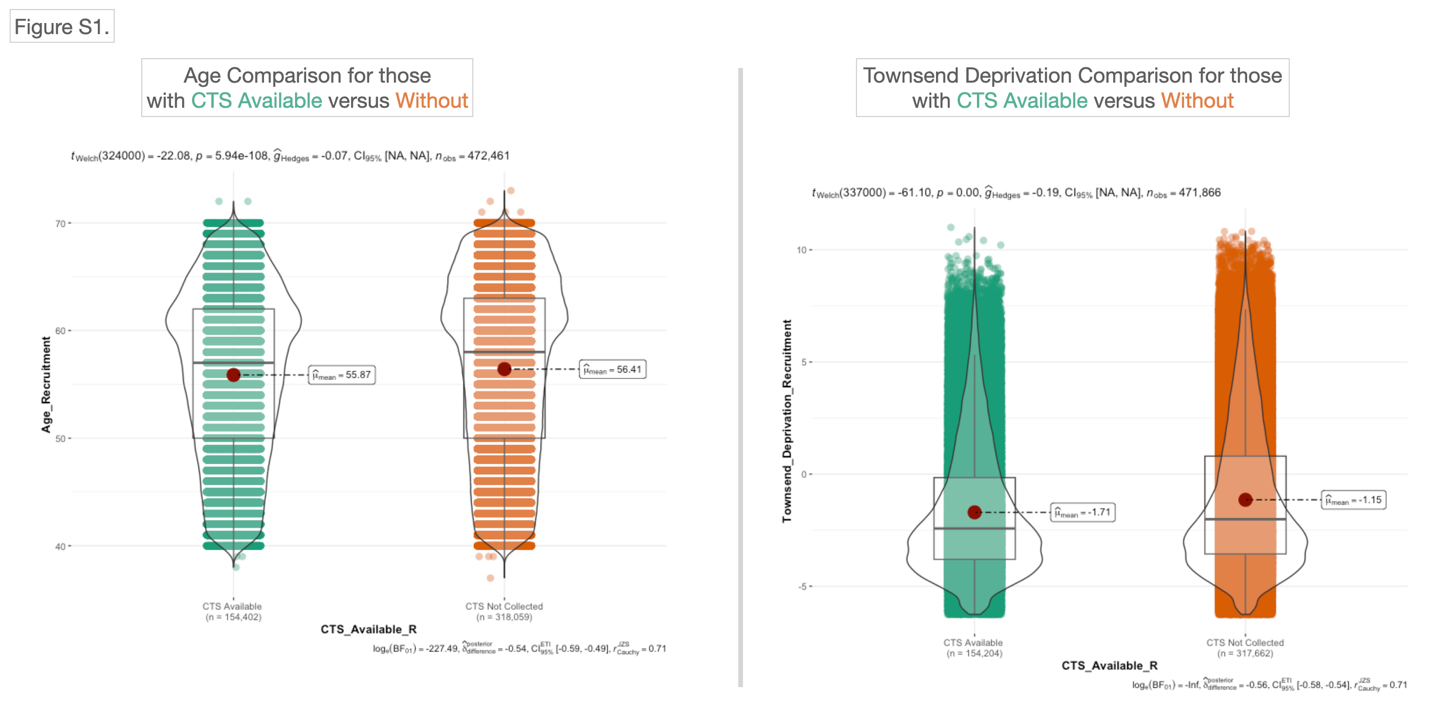

Caption: Participants who completed the Childhood Trauma Screener (CTS) were significantly younger at recruitment and more affluent compared to those who did not complete it. In the left panel, presence or absence of CTS is shown on the horizontal axis, and age at recruitment is shown on the vertical axis. In the right panel, presence or absence of CTS is shown on the horizontal axis, and Townsend Deprivation Index is shown on the vertical axis.

Figure S2.

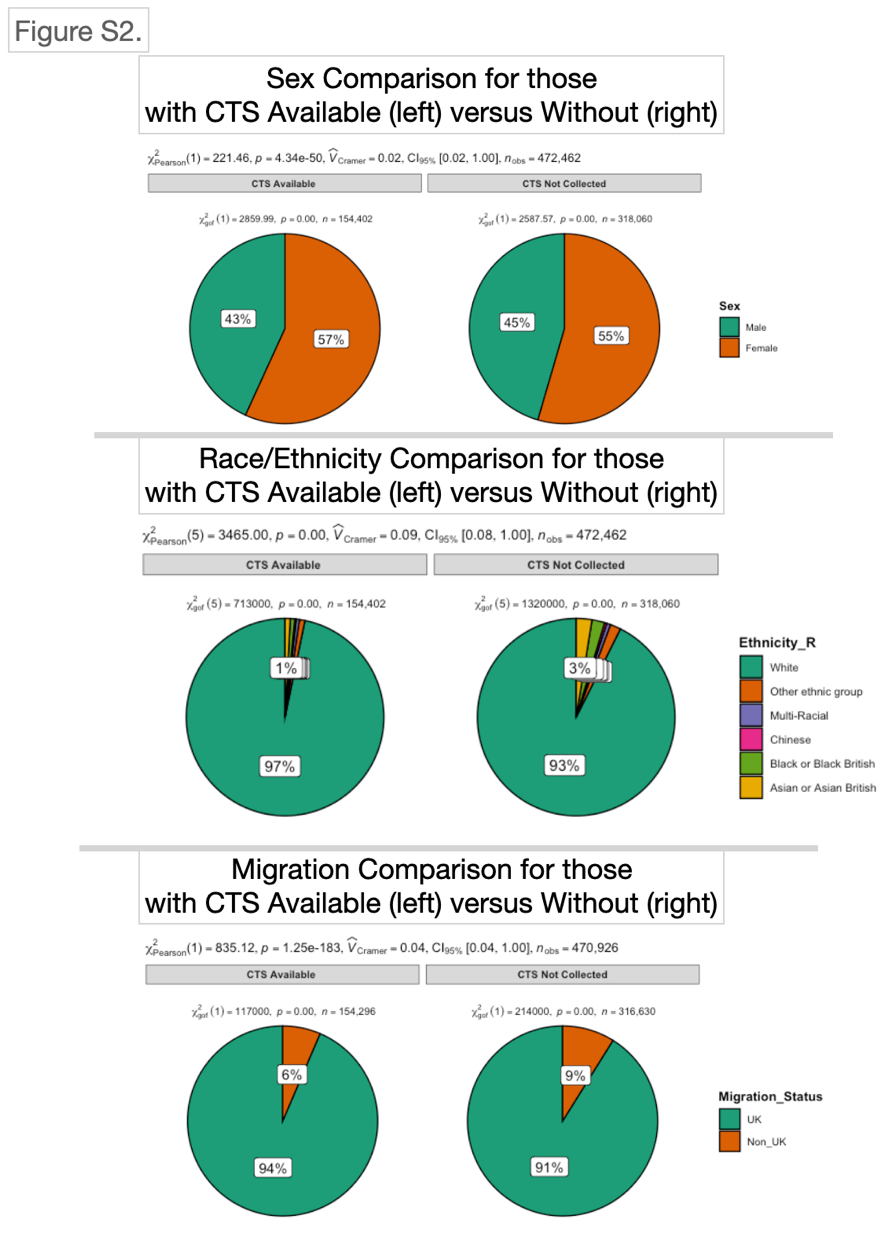

Caption: Completion of the Childhood Trauma Screener differed by sex, with more females completing it compared to males (top panel), had a higher percentage of white participants (middle panel), and had a lower percentage of Migrant participants (bottom panel). Each panel shows participants with (left side) or without (right side) CTS data. Colors in each panel indicate different sexes, ethnicities, or migration status.

Figure S3.

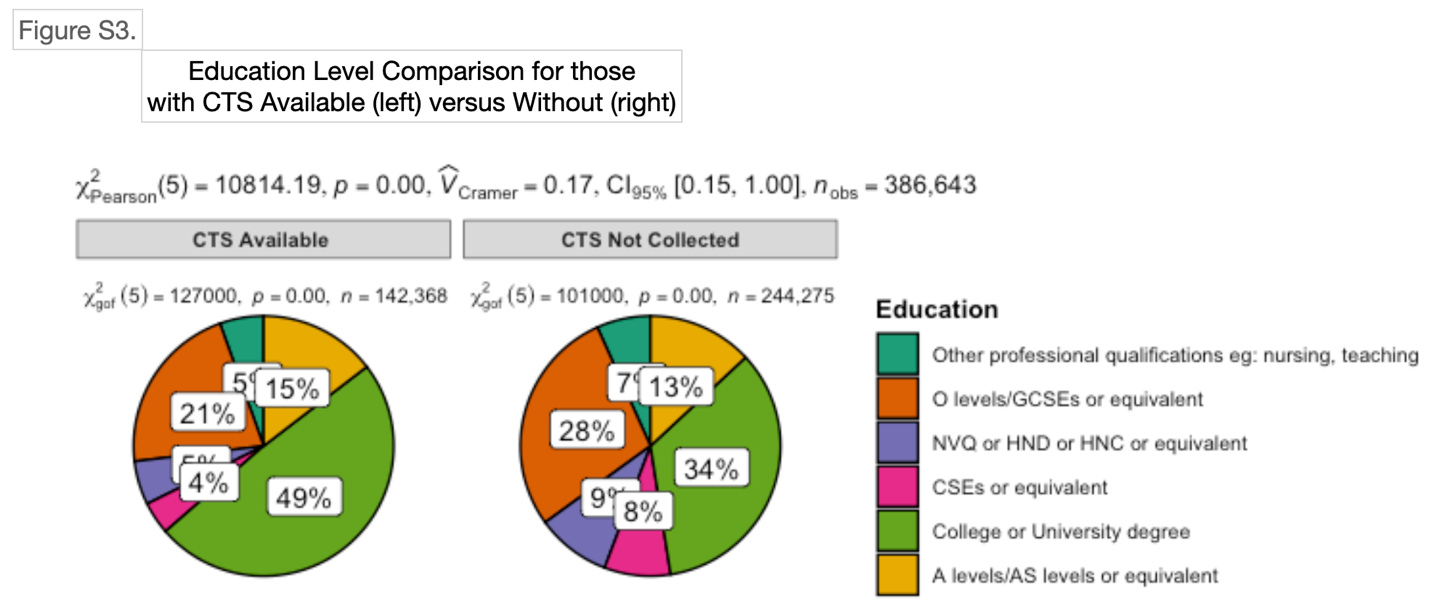

Caption: Participants who completed the Childhood Trauma Screener had higher levels of education. Participants with (left side) or without (right side) CTS data are shown here, with different colors representing education level.

Figure S4.

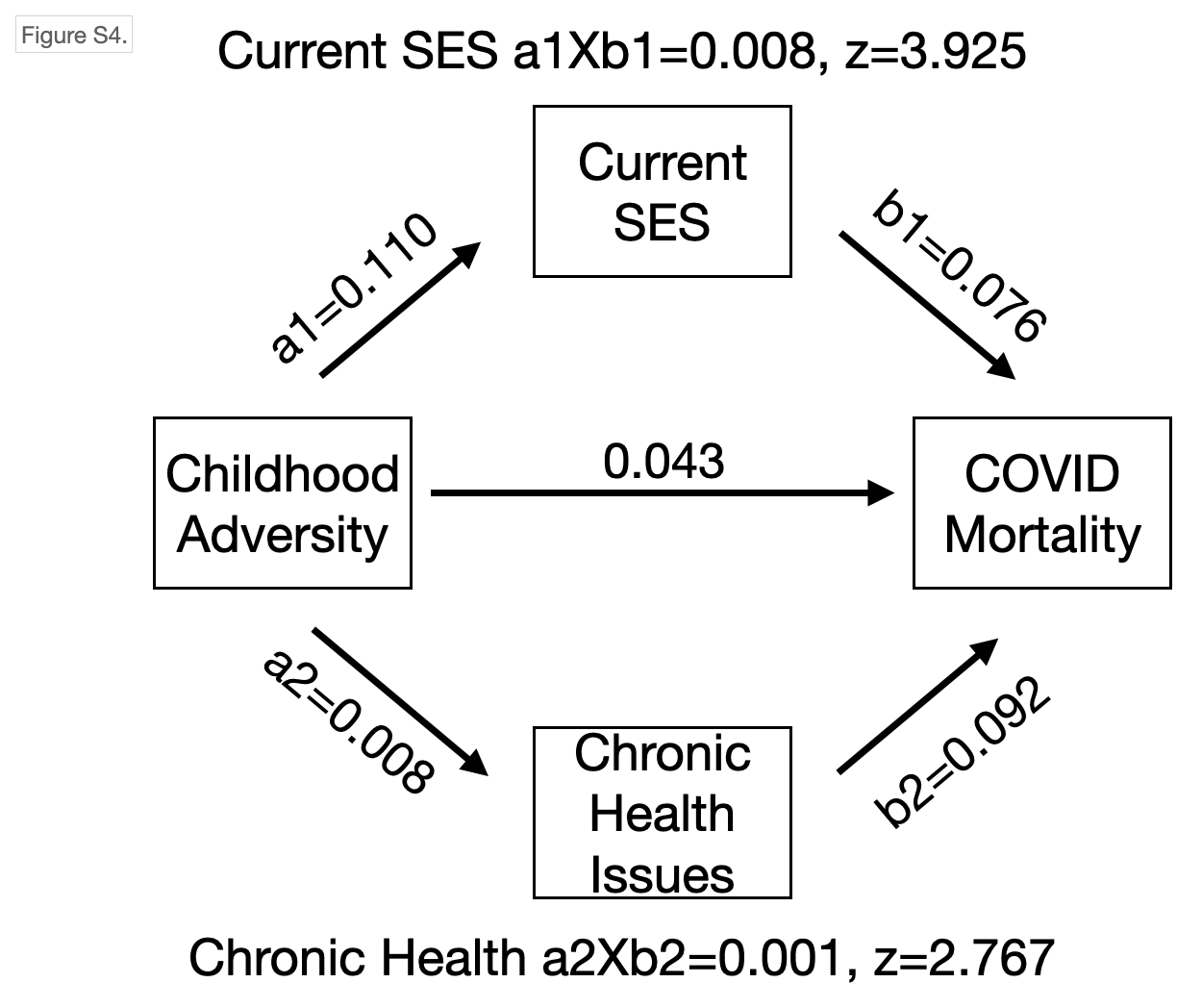

Caption: Indirect effects models showing current socioeconomic status and pre-existing health conditions partially (statistically) explain links between childhood adversity and COVID-19 mortality. Coefficients for each relation, along with z-statistics, are shown here.

Figure S5.

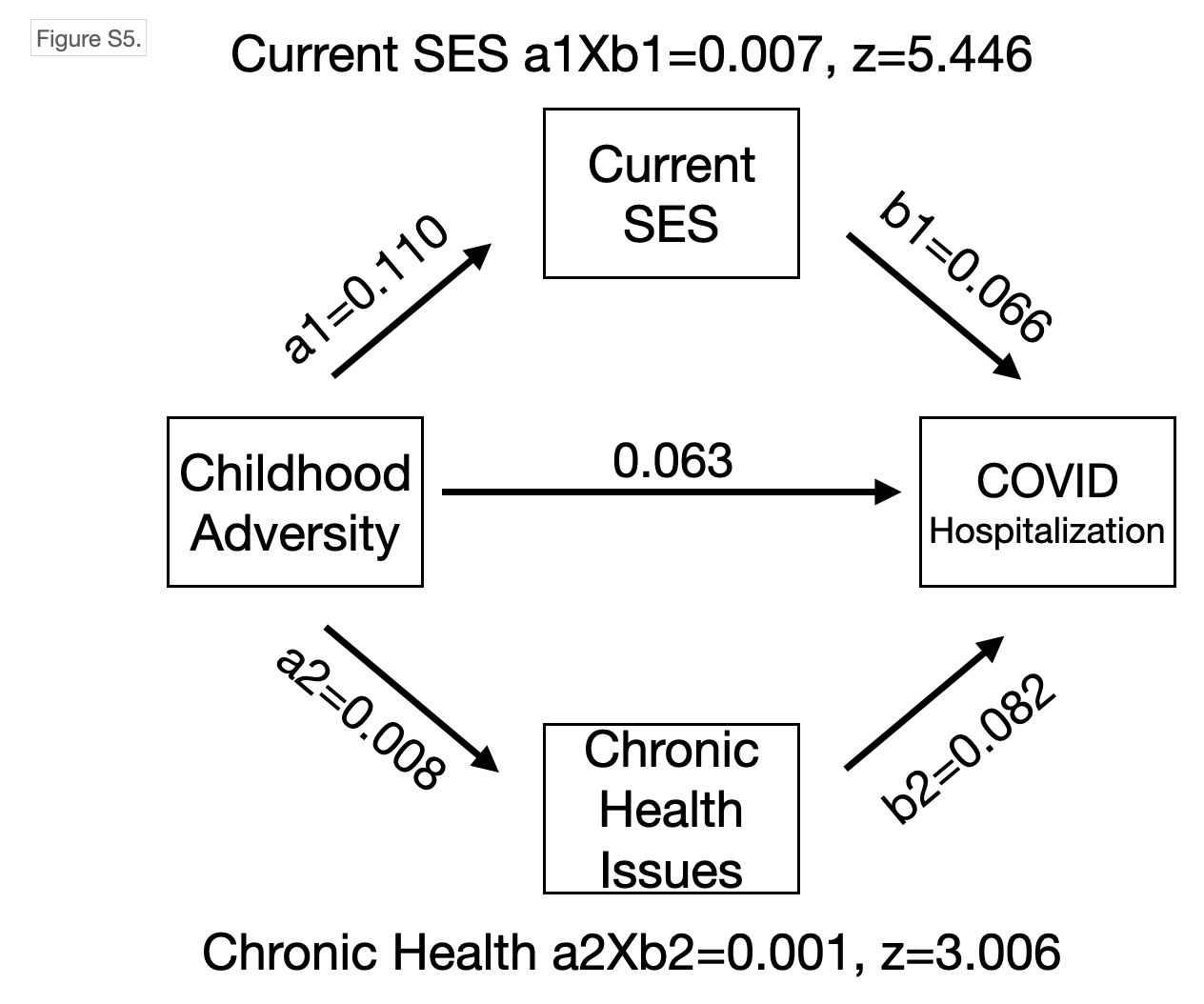

Caption: Indirect effects models showing current socioeconomic status and pre-existing health conditions partially (statistically) explain links between childhood adversity and COVID-19 hospitalization. Coefficients for each relation, along with z-statistics, are shown here.

Figure S6.

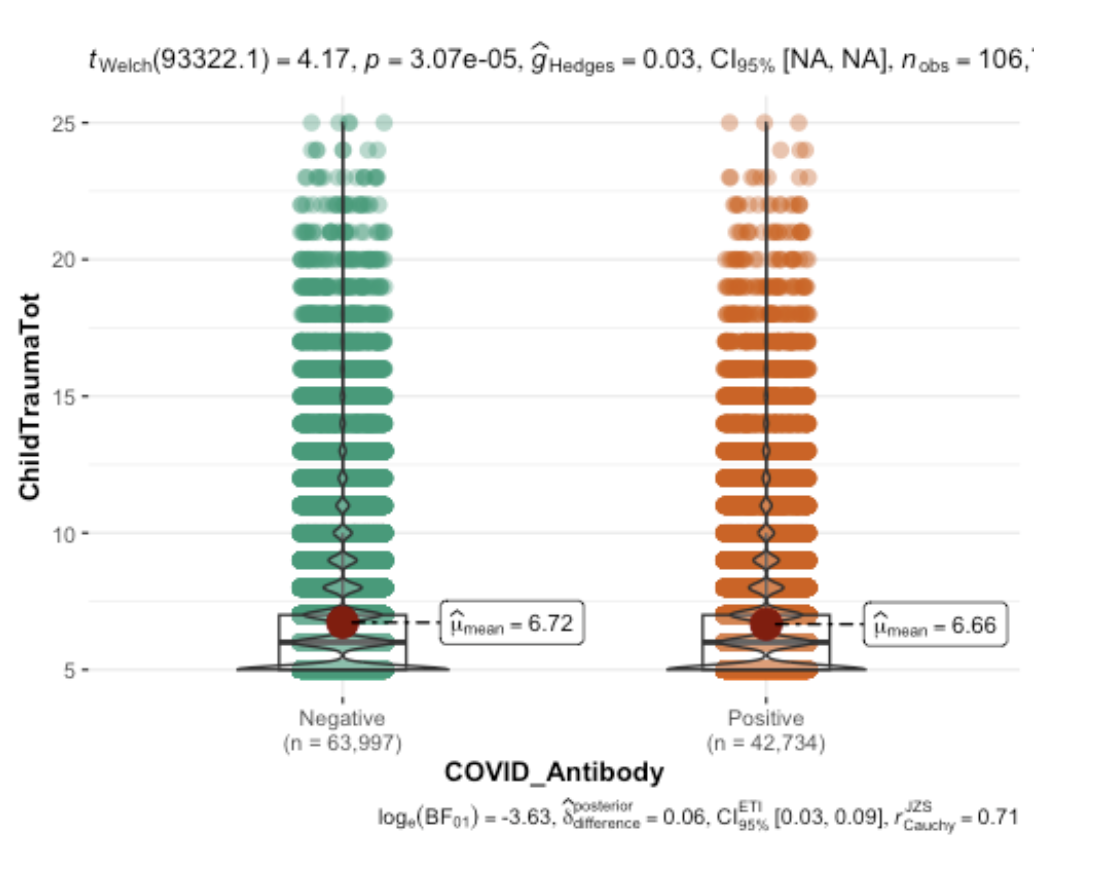

Caption: Childhood adversity was related to lower likelihood of having COVID-19 antibodies based on IgG/IgM antibody tests. Antibody test result is shown on the horizontal axis, while childhood adversity is shown on the vertical axis.
